# A national audit of pancreatic enzyme prescribing in pancreatic cancer from 2015 to 2023 in England using OpenSAFELY-TPP

**DOI:** 10.1101/2022.07.08.22277317

**Authors:** Agnieszka Lemanska, Colm Andrews, Louis Fisher, Ben Butler-Cole, Amir Mehrkar, Keith J Roberts, Ben Goldacre, Alex J Walker, The OpenSAFELY Collaborative, Brian MacKenna

## Abstract

**Objectives:** Cancer treatments were variably disrupted during the COVID-19 pandemic. UK guidelines recommend pancreatic enzyme replacement therapy (PERT) to all people with unresectable pancreatic cancer. The aim was to investigate the impact of COVID-19 on PERT prescribing to people with unresectable pancreatic cancer and to investigate the national and regional rates from January 2015 to January 2023.

**Data sources:** With the approval of NHS England, we conducted this study using 24 million electronic healthcare records of people within the OpenSAFELY-TPP research platform. There were 22,860 people diagnosed with pancreatic cancer in the study cohort. We visualised the trends over time and modelled the effect of COVID-19 with the interrupted time series analysis.

**Conclusions:** In contrast to many other treatments, prescribing of PERT was not affected during the pandemic. Overall, since 2015, the rates increased steadily over time by 1% every year. The national rates ranged from 41% in 2015 to 48% in early 2023. There was substantial regional variation with the highest rates of 50% to 60% in West Midlands.

**Implications for Nursing Practice:** In pancreatic cancer, if PERT is prescribed, it is usually initiated in hospitals by clinical nurse specialists and continued after discharge by primary care. At just under 50% in early 2023, the rates were still below the recommended 100% standard. More research is needed to understand barriers to prescribing of PERT and geographic variation to improve quality of care. Prior work relied on manual audits. With OpenSAFELY, we developed an automated audit allowing for regular updates.

## Background

Worldwide, there are around half a million new pancreatic cancer diagnoses every year (including 10,000 in the UK and 60,000 in the US).^1,2^ Currently, surgery is the only curative option, but with 85-90% of people diagnosed too late for surgical resection, pancreatic cancer has the lowest (9%) 5-year survival rate among all the cancers.^2^ Treatment options available to people with unresectable pancreatic cancer are palliative in nature and include symptom and pain management. Deficiency in the endo- and exocrine functions of the pancreas has implications for pancreatic cancer care. Diabetes presents commonly, and there are well established treatment pathways. Pancreatic exocrine failure, or insufficiency, is less well treated.^3-5^

There is clear evidence that untreated insufficient digestive enzyme production results in dramatic weight loss, malnutrition, impaired quality of life via gastrointestinal symptoms and, in pancreatic cancer, contributes to cachexia and frailty.^6,7^ In pancreatic cancer, pancreatic enzyme replacement therapy (PERT) can help to improve nutritional status and wellbeing and, as a result, the ability to tolerate the disease and treatment.^3,8-10^ A meta-analysis of the effect of PERT demonstrated a survival benefit of several months among patients with unresectable pancreatic cancer.^11^ This survival advantage is of similar magnitude to that associated with palliative chemotherapy.^11^ The first UK pancreatic cancer guidelines published by the National Institute for Health and Care Excellence (NICE) in February 2018, recommend PERT to people with pancreatic cancer.^12^ Subsequently, a Quality Standard (QS177 #4) was published by NICE in December 2018 to ensure that all adults with unresectable pancreatic cancer receive PERT.^13^

Despite the clear evidence of the benefits, as well as relatively low risks and few contraindications of PERT, the therapy remains underutilised in pancreatic cancer management.^3-5^ In addition, the recent healthcare disruption, associated with the COVID-19 pandemic, was shown to affect both primary care prescribing^14^ and cancer care.^15^ For people with pancreatic cancer, PERT is usually initiated in hospitals, with primary care continuing the prescription after discharge. Therefore, by analysing primary care prescribing we capture changes in the relevant clinical practice. Patients are more likely to receive PERT if a clinical nurse specialist or a dietician is involved in their hospital care.^16^ However, over 40% of patients do not receive a consultation with a dietician and over 20% do not see a clinical nurse specialist before they are discharged.^16^ Given that PERT prescribing was suboptimal prior to COVID-19, and that rationing of pancreatic cancer diagnostics and treatment has been reported during the pandemic across the UK, there is a concern that this patient population would be further disadvantaged during the pandemic.

We therefore set out to undertake a national audit of PERT prescribing in primary care from January 2015 to January 2023 to evaluate the impact of the COVID-19 pandemic and to investigate the national and regional prescribing rates over time. Previous work in this area, such as the largest (until this study) national audit undertaken by the RICOCHET Study Group in 2018,^16^ relied on manual audits of patient records. This is a resource intensive and limiting process (e.g., duration and frequency of audits). The methodological advance of OpenSAFELY offers an improved monitoring approach. OpenSAFELY facilitates the opportunity for transparent and reproducible automated audits which can be regularly updated with limited additional resources.

## Methods

### Study design

We used a cohort study design to assess the proportion of adults with unresectable pancreatic cancer receiving PERT from 1^st^ January 2015 to 31^st^ January 2023. The national target was 100%, as per the NICE (2018) Quality Standard.^13^

### Data Source

All data were linked, stored and analysed securely within the OpenSAFELY platform: opensafely.org. Primary care records managed by the GP software provider TPP were linked to Office of National Statistics (ONS) death data and to Secondary Uses Service (SUS) hospital procedures data through OpenSAFELY, a data analytics platform created by our team on behalf of NHS England to address urgent COVID-19 research questions (<u>opensafely.org</u>). OpenSAFELY provides a secure software interface allowing the analysis of pseudonymized primary care patient records from England in near real-time within the EHR vendor’s highly secure data centre, avoiding the need for large volumes of potentially disclosive pseudonymized patient data to be transferred off-site. This, in addition to other technical and organisational controls, minimises any risk of re-identification. The dataset was based on 24 million people currently registered with GP practices that use TPP’s SystmOne software (covering over 40% of England’s population). It included pseudonymized data such as coded diagnoses, medications and physiological parameters. It did not include free text data.

### Study population and study measures

Between 1^st^ January 2015 and 31^st^ January 2023, each month we identified individuals receiving PERT (numerator) among adults with unresectable pancreatic cancer (denominator). PERT was defined using a list of medications compiled from the British National Formulary (BNF, bnf.org) and coded using NHS Dictionary of Medicines and Devices, the nationally mandated NHS terminology. Individuals were classified each month as receiving PERT if they had a prescription between the 1^st^ day of that month and 61 days after. This was because 86% of prescriptions are issued for durations of one or two months.^17^ Date of pancreatic cancer diagnosis was defined as the first time that a clinical code for pancreatic cancer was entered in a primary care record. Adults with unresectable pancreatic cancer were defined by a presence of clinical codes for pancreatic cancer and an absence of OPCS-4 codes indicating a surgical resection procedure after pancreatic cancer diagnosis. The study flowchart explaining inclusion criteria is presented in Figure 1.

**Figure 1.**
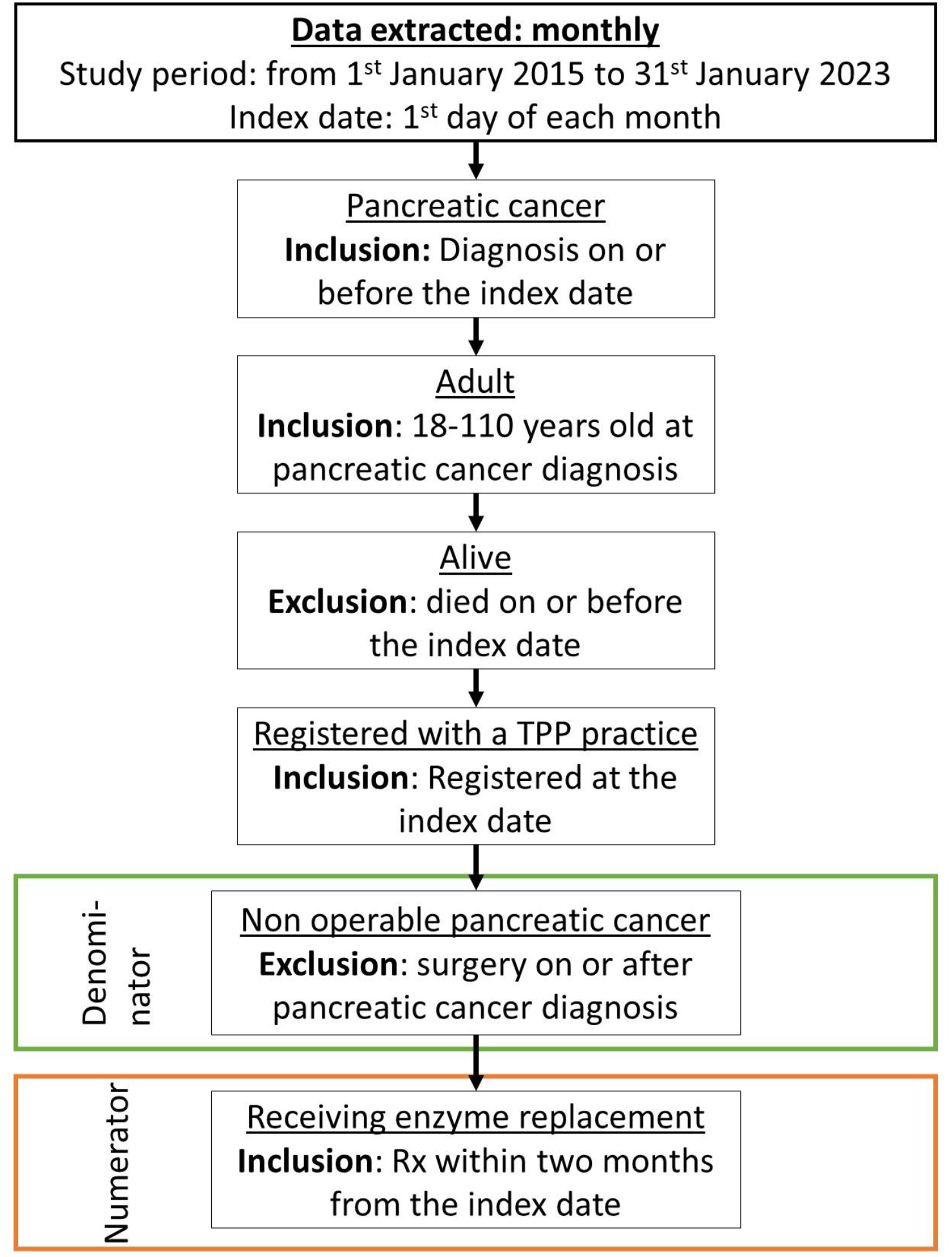
Study flowchart explaining inclusion and exclusion criteria. Each month, the rate was calculated of people receiving pancreatic enzyme replacement therapy (numerator) among people with unresectable pancreatic cancer (denominator).

### Statistical methods

The overall trends in prescribing, and by regions in England, were visualised over time. Monthly rates were presented as the proportion of individuals receiving PERT per 100 people with unresectable pancreatic cancer (%). Using the interrupted time series approach, generalised linear model (GLM) was fitted to the data to model the overall trend in prescribing over time and predict the expected rates in prescribing as if the pandemic had not occurred to model the effect of COVID-19. The time was included as a continuous variable to allow change in prescribing rates over time. The seasonality effect (month) was not accounted for. A variable that represented the COVID-19 period was included together with its time interaction term to allow varying slope. The 95% confidence intervals of the predicted values were used to estimate the significance of the difference between the predicted and observed values (to estimate the effect of the COVID-19 pandemic).

### Software and reproducibility

Data management was performed using Python 3.8 and analysis carried out using R 4.0.2. Software for data management and analysis is available online via github.com/opensafely/PaCa_Enzyme_Rx. Code lists are available via <u>opencodelists.org</u>.

### Patient and public involvement

OpenSAFELY has developed a publicly available website https://opensafely.org/ through which they invite any patient or member of the public to make contact regarding the broader OpenSAFELY project.

## Results

### Study population

In the study period, there were 22,860 people diagnosed with pancreatic cancer in the dataset. On average, there were 263 (±24 SD) pancreatic cancer diagnoses each month. As shown elsewhere, in this cohort of people, the COVID-19 pandemic did not affect the rates of pancreatic cancer diagnosis, but it negatively affected rates of surgical treatment ^18^. The mean age at pancreatic cancer diagnosis was 72 (±11 SD), 48% participants were females and 95% of participants (for which ethnicity data were recorded) were of White ethnicity.

### The effect of COVID-19

Figure 2 shows the rates in prescribing over time in England. Although, the average trends in the pandemic period are slightly lower than predicted, we concluded that overall, the COVID-19 pandemic did not affect PERT prescribing to people with unresectable pancreatic cancer. A clear dip in treatment rates, by about 3% (from 46% to 43%), was observed immediately at the start of the pandemic (from March to July 2020). This dip could be associated with the effect of COVID-19. However, the dip was small and transient, and the rates of prescribing recovered by September 2020 to rates that would be expected if the pandemic had not occurred. In addition, although the average trends throughout the pandemic period were lower than predicted (by about 1%), and this was statistically significant, this cannot be directly interpreted as the effect of the pandemic.

**Figure 2.**
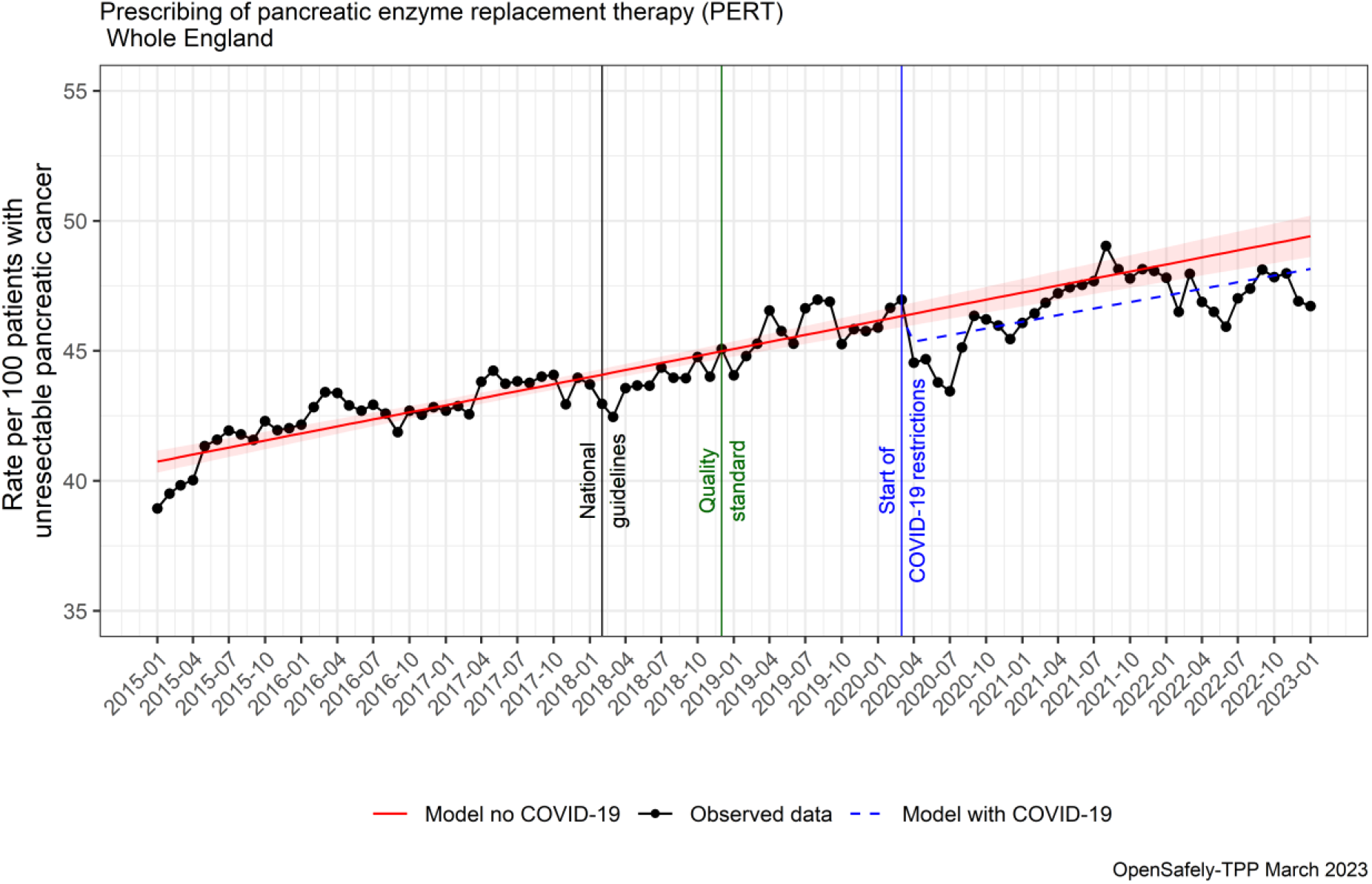
National rates of prescribing of pancreatic enzyme replacement therapy to people with unresectable pancreatic cancer in England between 1st January 2015 and 31^st^ January 2023. Vertical lines indicate points in time of interest to the study; black line, February 2018: publication of the National Institute for Health and Care Excellence (NICE) first UK national guidelines ^12^ recommending pancreatic enzyme replacement therapy to people with pancreatic cancer; green line, December 2018: publication of the NICE Quality Standard 4^13^ published to ensure that all adults with unresectable pancreatic cancer receive pancreatic enzyme supplementation; blue line, March 2020: start of the COVID-19-related national restrictions.

### National and regional trends

In view of the national target rate of 100%, the national trend increased gradually over time on average by 1% every year, ranging from 41% in 2015 to 48% by the end of 2022 (Figure 2). Figure 3 shows the rates in prescribing by regions in England. The highest rates, achieving values between 50% and 60% from 2018 onwards were in the West Midlands region. The lowest rates of between 20% to 30% were in the London region. However, because London is underrepresented in the dataset,^19^ these findings need to be interpreted with caution. The effect of COVID-19 was the most pronounced in the West Midlands region with a dip in rates by nearly 10% (from 50% to 40%) in July 2020. This recovered by September 2020 to levels higher than pre-pandemic.

**Figure 3.**
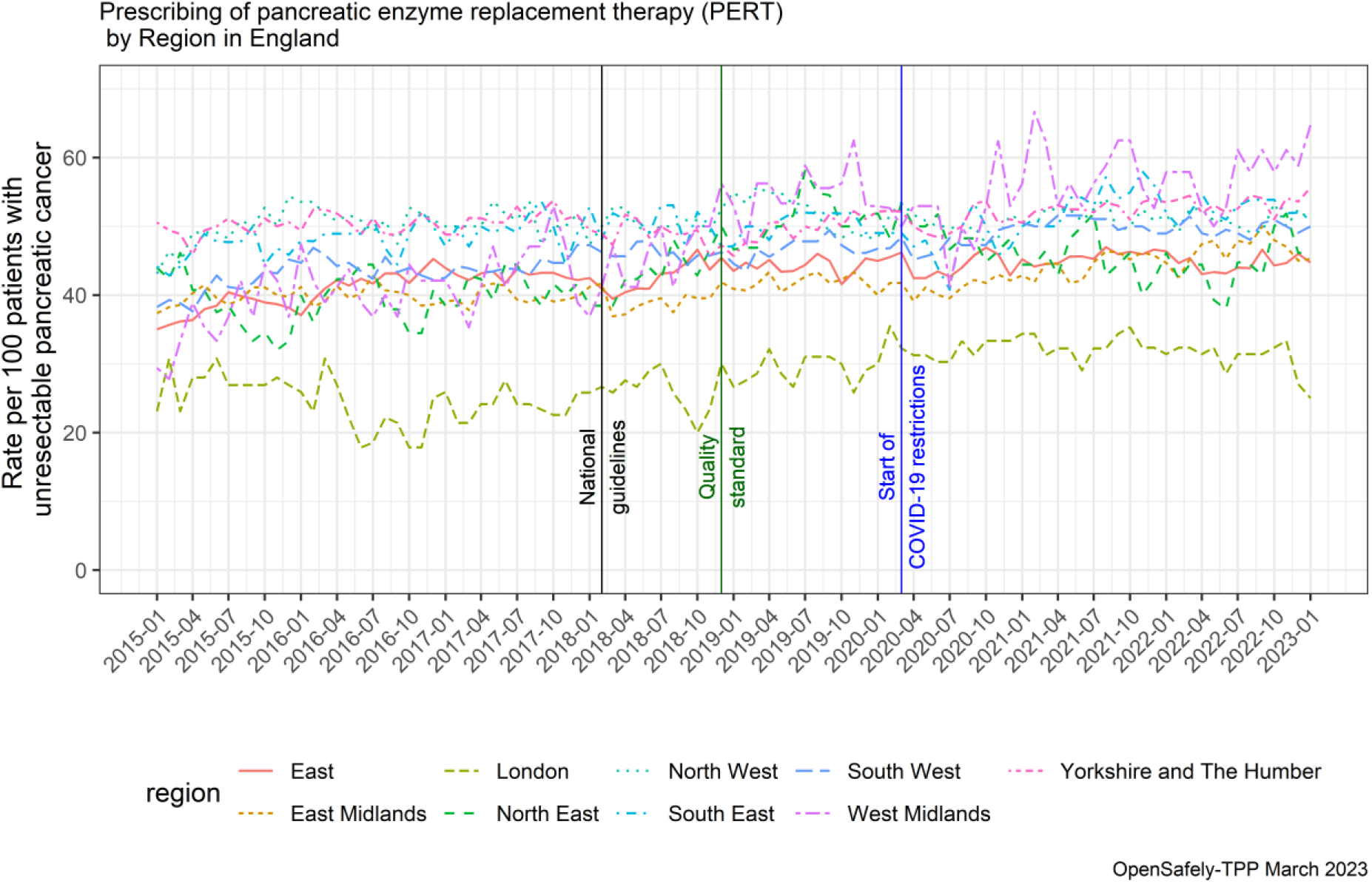
Regional rates of prescribing of pancreatic enzyme replacement therapy to people with unresectable pancreatic cancer in England between 1^st^ January 2015 and 31^st^ January 2023. Note the representativeness of the London region in the dataset is limited. Vertical lines indicate points in time of interest to the study; black line, February 2018: publication of the National Institute for Health and Care Excellence (NICE) first UK national guidelines^12^ recommending pancreatic enzyme replacement therapy to people with pancreatic cancer; green line, December 2018: publication of the NICE Quality Standard 4^13^ published to ensure that all adults with unresectable pancreatic cancer receive pancreatic enzyme supplementation; blue line, March 2020: beginning of the COVID-19-related national restrictions.

## Discussion

Overall, there was no change in prescribing of PERT to people with unresectable pancreatic cancer during the COVID-19 pandemic. Reassuringly, despite the challenges of the COVID-19 pandemic, clinicians ensured continuity of care in this area. We did observe a small and temporary decrease in prescribing rates from March to July 2020, which recovered fully by September 2020 to rates that would be expected if the pandemic had not happened. The delivery of cancer services was severely impacted by the COVID-19 pandemic. Effects upon cancer surgery,^20,21^ chemotherapy^22^ and cancer research^23^ have been well described. In pancreatic cancer, centres have described impact upon the entire pathway, including diagnostic, staging as well as treatment delivery.^24^ In contrast, in this study we show that prescribing of PERT to people with unresectable pancreatic cancer was not affected during the COVID-19 pandemic.

Importantly, with the target prescribing rate of 100% (all adults with unresectable pancreatic cancer are to be prescribed PERT),^13^ we found that despite the clear benefits, PERT remained under-prescribed in this group of patients. In pancreatic cancer, PERT is usually initiated in hospitals and primary care continues the prescription after discharge. The RICOCHET audit demonstrated that patients were more likely to receive PERT if clinical nurse specialists or dieticians were involved in patients’ care and suggested that services should ensure patients have contact with these clinicians during their hospital care.^16^ Clinical nurse specialists have an important role in educating patients regarding the correct administration of PERT and continuation of PERT after discharge and should ensure dissemination of best practice to primary care.

A previous national UK study, undertaken between 2001 and 2015, demonstrated an overall prescribing rate of PERT to people with pancreatic cancer at 21%.^3^ A regional UK study observed a prescribing rate of 22% between 2010 and 2012.^4^ More recently, the national RICOCHET audit conducted April to August 2018, six months after the introduction of the national pancreatic cancer guidelines, demonstrated rates of 74% for patients with resectable and 45% with unresectable disease.^16^ The current study demonstrated rates consistent with the RICOCHET audit. However an important finding is that although increasing over time, at just under 50% in 2022, the rates were still below the recommended 100% quality standard.^13^

### Strengths and limitations

The key strength of this population-based study is the large size (40% of the population in England) and completeness of the underlying raw EHR data in OpenSAFELY.^25^ In addition, because the OpenSAFELY platform provides analysis directly on the servers of the healthcare records administrator (TPP), we access near real-time data while prioritising patient privacy (no need for data transfers). The national audit undertaken by the RICOCHET Study Group,^16^ relied on manual audits of patient records by many local teams in the UK, following by compilation of the results centrally. By contrast, the automated audit method that we developed, enables frequent and cost-effective re-execution of the analysis at regular audit cycles. In addition, because with OpenSAFELY we are able to access the full set of patients’ records, we provided a longitudinal view of trends in prescribing since 2015. This allowed us to investigate the effect of the COVID-19 pandemic. All analytics software and code lists are shared openly and are available for inspection and re-use, providing opportunity for reproduction of this report to support recovery from the COVID-19 pandemic and reducing duplicative efforts.

We also note some limitations. We note that our data will only include prescriptions issued in primary care, it is possible that some hospitals supply the medicines directly to patients when they attend the hospital. We have written extensively^26,27^ on the availability of hospital prescriptions and NHS Digital have recently made some of this hospital medication data available for a small subset of hospitals ^28^. We will seek to incorporate this in any future work. In addition to regional variation, more research is needed to investigate different groups of patients. The pandemic exacerbated healthcare inequalities and future work is needed to investigate PERT prescribing in different socio-demographic groups. We also note that pancreatic cancer was defined by coding in primary care, rather than via linkage with cancer registry, the gold standard for cancer diagnoses.

### Policy Implications and future research

Across the UK, pancreatic cancer care has been centralised since 2001, largely due to the need to reduce perioperative mortality. This, however, created a potential disconnect between specialist and non-specialist sites. Patients with unresectable pancreatic cancer are typically treated at their local hospital, which for most patients will not be a specialist site and will receive regular repeat prescriptions from their general practitioner. The RICOCHET audit of pancreatic cancer and PERT prescribing demonstrated that rates of PERT prescribing were highest within specialist sites, regardless of tumour stage and treatment intent.^16^ RICOCHET also demonstrated that high prescribing rates where shared within hospitals (i.e., if there was high prescribing in resectable cancer, there was high prescribing in unresectable cancer and vice versa), but not across each local network which consisted of the central specialist surgical unit and their referring non-specialist sites (i.e., prescribing rates in each specialist centre did not correlate with those within each centre’s network of referring non-specialist sites). The geographic variation observed in the present study contrasts with that data. Given that RICOCHET was a snapshot of a 3-month cohort of patients, the present study is much better placed to observe regional variation in practice. Understanding why regions have differing rates of prescribing could help overcome barriers to PERT prescribing.

Additionally, qualitative research, beyond the scope of this study, is needed to understand barriers to the implementation of NICE guidelines and quality standards.

More broadly, the implications of this study for data usage to improve NHS services are very substantial. Previously, practical and privacy challenges around accessing GP data meant that the largest study to date in this area, RICOCHET, relied on manual audits by many local teams at a single point in time. This manual approach imposes a substantial resource burden on local teams collecting data as well as being hard to reproduce on an ongoing basis. Using the OpenSAFELY framework we were able to execute a single analysis in OpenSAFELY-TPP for 40% of the population in near real-time, whilst leaving the data in situ, preserving trust. OpenSAFELY was the single most trusted COVID-19 data project in a rigorous Citizen’s Jury sponsored by the NHS and the National Data Guardian.^29^ We can extend this analysis to OpenSAFELY-EMIS, increasing coverage to 99% of English general practices, as well as providing fine grained demographic (e.g., ethnicity) or clinical sub-populations. OpenSAFELY tools can facilitate cost-effective, ongoing and near real-time audits and feedback to NHS organisations. This presents an important opportunity in the context of rapidly evolving pressures on the health service during the COVID-19 recovery period.

## Conclusions

The COVID-19 pandemic did not affect PERT prescribing in unresectable pancreatic cancer. However, despite national guidelines, under-prescribing of PERT continues and has improved only marginally since their publication. This could be an important missed opportunity to reduce morbidity for patients. The research into the effect of COVID-19 as well as barriers to prescribing of PERT and geographic variation is urgently needed to improve quality of care.

## Data Availability

Detailed pseudonymised patient data are potentially re-identifiable and therefore not shared. The process for external users to request access to data via the OpenSAFELY platform is described on opensafely.org.

https://www.opensafely.org/

## List of abbreviations

COVID-19: Coronavirus disease 2019
EHR: Electronic healthcare records
GP: General practice
NHS: National Health Service
PERT: Pancreatic enzyme replacement therapy
RICOCHET: Receipt of curative resection or palliative care for hepatopancreaticobiliary tumours

## Statements

## Acknowledgements

We are very grateful for all the support received from the TPP Technical Operations team throughout this work, and for generous assistance from the information governance and database teams at NHS England and the NHS England Transformation Directorate.

Membership of the OpenSAFELY Collaborative: Alex J. Walker, Brian MacKenna, Peter Inglesby, Christopher T. Rentsch, Helen J. Curtis, Caroline E. Morton, Jessica Morley, Amir Mehrkar, Seb Bacon, George Hickman, Chris Bates, Richard Croker, David Evans, Tom Ward, Jonathan Cockburn, Simon Davy, Krishnan Bhaskaran, Anna Schultze, Elizabeth J. Williamson, William J. Hulme, Helen I. McDonald, Laurie Tomlinson, Rohini Mathur, Rosalind M. Eggo, Kevin Wing, Angel Y. S. Wong, Harriet Forbes, John Tazare, John Parry, Frank Hester, Sam Harper, Ian J. Douglas, Stephen J. W. Evans, Liam Smeeth & Ben Goldacre

## Contributions

The OpenSAFELY Collaborative built the data infrastructure used in this study; AL, AM, AW and BMK contributed to the conception and planning of the study; AL, CA, LF, BBC, AW and BMK conducted the analysis; AL, CA, LF, KJR, BG, AW and BMK contributed to the interpretation of the results; AL, KJR and BMK written the first draft of the manuscript; all authors contributed to the subsequent revisions and approved the final version.

## Information governance and ethical approval

NHS England is the data controller for OpenSAFELY-TPP; TPP is the data processor; all study authors using OpenSAFELY have the approval of NHS England. This implementation of OpenSAFELY is hosted within the TPP environment which is accredited to the ISO 27001 information security standard and is NHS IG Toolkit compliant.^30^

Patient data has been pseudonymised for analysis and linkage using industry standard cryptographic hashing techniques; all pseudonymised datasets transmitted for linkage onto OpenSAFELY are encrypted; access to the platform is via a virtual private network (VPN) connection, restricted to a small group of researchers; the researchers hold contracts with NHS England and only access the platform to initiate database queries and statistical models; all database activity is logged; only aggregate statistical outputs leave the platform environment following best practice for anonymisation of results such as statistical disclosure control for low cell counts.^31^

The OpenSAFELY research platform adheres to the obligations of the UK General Data Protection Regulation (GDPR) and the Data Protection Act 2018. In March 2020, the Secretary of State for Health and Social Care used powers under the UK Health Service (Control of Patient Information) Regulations 2002 (COPI) to require organisations to process confidential patient information for the purposes of protecting public health, providing healthcare services to the public and monitoring and managing the COVID-19 outbreak and incidents of exposure; this sets aside the requirement for patient consent.^32^ This was extended in November 2022 for the NHS England OpenSAFELY COVID-19 research platform.^33^ In some cases of data sharing, the common law duty of confidence is met using, for example, patient consent or support from the Health Research Authority Confidentiality Advisory Group.^34^

Taken together, these provide the legal bases to link patient datasets on the OpenSAFELY platform. GP practices, from which the primary care data are obtained, are required to share relevant health information to support the public health response to the pandemic, and have been informed of the OpenSAFELY analytics platform.

The study was approved by the Health Research Authority (Research Ethics Committee reference 20/LO/0651) and the London School of Hygiene and Tropical Medicine (London, UK) Ethics Board (reference 21863).

## Data access and verification

Access to the underlying identifiable and potentially re-identifiable pseudonymised electronic health record data is tightly governed by various legislative and regulatory frameworks, and restricted by best practice. The data in OpenSAFELY is drawn from General Practice data across England where TPP is the data processor. TPP developers initiate an automated process to create pseudonymised records in the core OpenSAFELY database, which are copies of key structured data tables in the identifiable records. These pseudonymised records are linked onto key external data resources that have also been pseudonymised via SHA-512 one-way hashing of NHS numbers using a shared salt. Bennett Institute for Applied Data Science developers and PIs holding contracts with NHS England have access to the OpenSAFELY pseudonymised data tables as needed to develop the OpenSAFELY tools. These tools in turn enable researchers with OpenSAFELY data access agreements to write and execute code for data management and data analysis without direct access to the underlying raw pseudonymised patient data, and to review the outputs of this code. All code for the full data management pipeline - from raw data to completed results for this analysis - and for the OpenSAFELY platform as a whole is available for review at github.com/OpenSAFELY. The data management and analysis code for this paper was led by AL and contributed to by CA.

## References

1. Sung H, Ferlay J, Siegel RL, et al. Global Cancer Statistics 2020: GLOBOCAN Estimates of Incidence and Mortality Worldwide for 36 Cancers in 185 Countries. CA: A Cancer Journal for Clinicians. 2021;71(3):209–249. doi:https://doi.org/10.3322/caac.21660

2. Siegel RL, Miller KD, Jemal A. Cancer statistics, 2020. CA Cancer J Clin. Jan 2020;70(1):7–30. doi:10.3322/caac.21590

3. Roberts KJ, Bannister CA, Schrem H. Enzyme replacement improves survival among patients with pancreatic cancer: Results of a population based study. Pancreatology : official journal of the International Association of Pancreatology (IAP) [et al]. Jan 2019;19(1):114–121. doi:10.1016/j.pan.2018.10.010

4. Landers A, Muircroft W, Brown H. Pancreatic enzyme replacement therapy (PERT) for malabsorption in patients with metastatic pancreatic cancer. BMJ Supportive & Palliative Care. 2016;6(1):75–79. doi:10.1136/bmjspcare-2014-000694

5. Sikkens EC, Cahen DL, van Eijck C, Kuipers EJ, Bruno MJ. The daily practice of pancreatic enzyme replacement therapy after pancreatic surgery: a northern European survey: enzyme replacement after surgery. J Gastrointest Surg. Aug 2012;16(8):1487–92. doi:10.1007/s11605-012-1927-1

6. Armstrong T, Walters E, Varshney S, Johnson CD. Deficiencies of micronutrients, altered bowel function, and quality of life during late follow-up after pancreaticoduodenectomy for malignancy. Pancreatology : official journal of the International Association of Pancreatology (IAP) [et al]. 2002;2(6):528–34. doi:10.1159/000066095

7. Wigmore SJ, Plester CE, Richardson RA, Fearon KC. Changes in nutritional status associated with unresectable pancreatic cancer. Br J Cancer. 1997;75(1):106–9. doi:10.1038/bjc.1997.17

8. Pezzilli R, Caccialanza R, Capurso G, Brunetti O, Milella M, Falconi M. Pancreatic Enzyme Replacement Therapy in Pancreatic Cancer. Cancers. Jan 22 2020;12(2)doi:10.3390/cancers12020275

9. Guidelines for the management of patients with pancreatic cancer periampullary and ampullary carcinomas. Gut. 2005;54(uppl 5):v1–v16. doi:10.1136/gut.2004.057059

10. Phillips ME, Hopper AD, Leeds JS, et al. Consensus for the management of pancreatic exocrine insufficiency: UK practical guidelines. BMJ Open Gastroenterology. 2021;8(1):e000643. doi:10.1136/bmjgast-2021-000643

11. Iglesia D, Avci B, Kiriukova M, et al. Pancreatic exocrine insufficiency and pancreatic enzyme replacement therapy in patients with advanced pancreatic cancer: A systematic review and meta-analysis. United European Gastroenterol J. Nov 2020;8(9):1115–1125. doi:10.1177/2050640620938987

12. The National Institute for Health and Care Excellence (NICE) 2018. Pancreatic cancer in adults: diagnosis and management. NICE guideline [NG85] https://www.nice.org.uk/guidance/ng85 (accessed February 2022).

13. The National Institute for Health and Care Excellence (NICE) 2018. Pancreatic cancer Quality standard [QS177]. https://www.nice.org.uk/guidance/qs177/chapter/Quality-statement-4-Pancreatic-enzyme-replacement-therapy (accessed April 2023).

14. Mercier G, Arquizan C, Roubille F. Understanding the effects of COVID-19 on health care and systems. The Lancet Public Health. 2020;5(10):e524. doi:10.1016/S2468-2667(20)30213-9

15. Richards M, Anderson M, Carter P, Ebert BL, Mossialos E. The impact of the COVID-19 pandemic on cancer care. Nature Cancer. 2020/06/01 2020;1(6):565–567. doi:10.1038/s43018-020-0074-y

16. RICOCHET Study Group on behalf of the West Midlands Research Collaborative. Pancreatic enzyme replacement therapy in patients with pancreatic cancer: A national prospective study. Pancreatology : official journal of the International Association of Pancreatology (IAP) [et al]. May 25 2021;doi:10.1016/j.pan.2021.05.299

17. The Centre for Evidence-Based Medicine (CEBM) 2020. Should we prescribe longer repeat prescriptions for patients with long-term conditions during a pandemic? https://www.cebm.net/covid-19/should-we-prescribe-longer-repeat-prescriptions-for-patients-with-long-term-conditions-during-a-pandemic/ (accessed May 2022).

18. Lemanska A, Andrews C, Fisher L, et al. Healthcare in England was affected by the COVID-19 pandemic across the pancreatic cancer pathway: a cohort study using OpenSAFELY-TPP. medRxiv. 2022:2022.12.02.22283026. doi:10.1101/2022.12.02.22283026

19. Andrews C, Schultze A, Curtis H, et al. OpenSAFELY: Representativeness of electronic health record platform OpenSAFELY-TPP data compared to the population of England. Wellcome Open Res. 2022;7:191. doi:10.12688/wellcomeopenres.18010.1

20. Glasbey J, Ademuyiwa A, Adisa A, et al. Effect of COVID-19 pandemic lockdowns on planned cancer surgery for 15 tumour types in 61 countries: an international, prospective, cohort study. The Lancet Oncology. 2021;22(11):1507–1517. doi:10.1016/S1470-2045(21)00493-9

21. Nepogodiev D, Abbott TEF, Ademuyiwa AO, et al. Projecting COVID-19 disruption to elective surgery. The Lancet. 2022;399(10321):233–234. doi:10.1016/S0140-6736(21)02836-1

22. Morris EJA, Goldacre R, Spata E, et al. Impact of the COVID-19 pandemic on the detection and management of colorectal cancer in England: a population-based study. The Lancet Gastroenterology & Hepatology. 2021;6(3):199–208. doi:10.1016/S2468-1253(21)00005-4

23. Bakouny Z, Labaki C, Bhalla S, et al. Oncology clinical trial disruption during the COVID-19 pandemic: a COVID-19 and cancer outcomes study. Ann Oncol. Jun 10 2022; doi:10.1016/j.annonc.2022.04.071

24. McKay SC, Pathak S, Wilkin RJW, et al. Impact of SARS-CoV-2 pandemic on pancreatic cancer services and treatment pathways: United Kingdom experience. HPB (Oxford). Nov 2021;23(11):1656–1665. doi:10.1016/j.hpb.2021.03.003

25. Andrews CD, Schultze A, Curtis HJ, et al. OpenSAFELY: Representativeness of Electronic Health Record platform OpenSAFELY-TPP data compared to the population of England. medRxiv. 2022:2022.06.23.22276802. doi:10.1101/2022.06.23.22276802

26. Goldacre B, MacKenna B. The NHS deserves better use of hospital medicines data. BMJ. 2020;370:m2607. doi:10.1136/bmj.m2607

27. Rowan A, Bates C, Hulme W, et al. A comprehensive high cost drugs dataset from the NHS in England - An OpenSAFELY-TPP Short Data Report. Wellcome Open Res. 2021;6:360. doi:10.12688/wellcomeopenres.17360.1

28. NHS Digital, 2020, Electronic Prescribing and Medicines Administration (EPMA) Data. https://digital.nhs.uk/about-nhs-digital/corporate-information-and-documents/directions-and-data-provision-notices/data-provision-notices-dpns/electronic-prescribing-and-medicines-administration-data (accessed June 2022).

29. Oswald M., Laverty L., 2021, Juries’ Report: Data Sharing in a Pandemic: Three Citizens’ Juries. https://arc-gm.nihr.ac.uk/media/Resources/ARC/Digital%20Health/Citizen%20Juries/New%2012621_NIHR_Juries_Report_WEB.pdf (accessed June 2022).

30. Data Security and Protection Toolkit - NHS Digital. NHS Digital. https://digital.nhs.uk/data-and-information/looking-after-information/data-security-and-information-governance/data-security-and-protection-toolkit (accessed 30 Apr 2020).

31. ISB1523: Anonymisation Standard for Publishing Health and Social Care Data - NHS Digital. NHS Digital. https://digital.nhs.uk/data-and-information/information-standards/information-standards-and-data-collections-including-extractions/publications-and-notifications/standards-and-collections/isb1523-anonymisation-standard-for-publishing-health-and-social-care-data (accessed 30Apr 2020).

32. Secretary of State for Health and Social Care - UK Government. Coronavirus (COVID-19): notification to organisations to share information. 2020. https://web.archive.org/web/20200421171727/https://www.gov.uk/government/publications/coronavirus-covid-19-notification-of-data-controllers-to-share-information (accessed June 2022).

33. Secretary of State for Health and Social Care - UK Government. Coronavirus (COVID-19): notification to organisations to share information. 2022. https://www.gov.uk/government/publications/coronavirus-covid-19-notification-to-organisations-to-share-information/coronavirus-covid-19-notice-under-regulation-34-of-the-health-service-control-of-patient-information-regulations-2002 (accessed April 2023).

34. Confidentiality Advisory Group. Health Research Authority. https://www.hra.nhs.uk/about-us/committees-and-services/confidentiality-advisory-group/ (accessed April 2023).

